# Development of *CuidaTEXT*: a text message intervention to support Latino dementia family caregivers

**DOI:** 10.1101/2021.12.06.21267369

**Authors:** Jaime Perales-Puchalt, Mariola Acosta-Rullán, Mariana Ramírez-Mantilla, Paul Espinoza-Kissell, Eric D. Vidoni, Michelle Niedens, Edward F. Ellerbeck, Ladson Hinton, Linda Loera, A. Susana Ramírez, Esther Lara, Amber Watts, Kristine Williams, Jason Resendez, Jeffrey Burns

**Author notes:** Corresponding Author: Jaime Perales Puchalt, PhD, MPH, KU Alzheimer’s Disease Research Center, 4350 Shawnee Mission Parkway, Fairway, KS 66205. Note: Mariola Acosta-Rullán is now at the University of Southern California, Los Angeles, CA; studying a PhD program. Paul Espinoza-Kissell is now at the University of Minnesota, Minneapolis, MN; studying a PhD program.

## Abstract

**Objectives:** To describe the development of *CuidaTEXT:* a tailored text message intervention to support Latino dementia family caregivers.

**Methods:** *CuidaTEXT* is informed by the Stress Process Framework and Social Cognitive Theory. We developed and refined *CuidaTEXT* using a mixed-method approach that included thematic analysis and descriptive statistics. We followed six user-centered design stages, including the selection of design principles, software vendor collaboration, evidence-based foundation, caregiver and research/clinical advisory board guidance, sketching and prototyping, and usability testing among five Latino caregivers.

**Results:** *CuidaTEXT* is a bilingual 6-month long intervention tailored to caregiver needs that includes: 1) 1-3 daily automatic messages (n=244) about logistics, dementia education, self-care, social support, end-of-life, care of the person with dementia, behavioral symptoms and problem-solving strategies; 2) 783 keyword-driven text messages for further help with the above topics; 3) live chat interaction with a coach for further help; 4) a 19-page reference booklet summarizing the purpose and functions of the intervention. *CuidaTEXT’s* prototype scored 97 out of 100 in the System Usability Scale.

**Conclusions:** *CuidaTEXT’s* prototype demonstrated high usability among Latino caregivers. *CuidaTEXT’s* feasibility is ready to be tested.

**Clinical Implications:** *CuidaTEXT’s* usability and its potential for widespread implementation holds promise in supporting Latino caregivers.

## Introduction

Family caregiving for individuals with dementia (IWDs) has a serious emotional, physical, and financial toll (Alzheimer’s Association, 2016; Bertrand et al., 2006; Friedman et al., 2015; Kasper et al., 2015; Moon et al., 2016; National Academies of Sciences & Medicine, 2016; Ory et al., 1999; Pinquart & Sörensen, 2005). Most IWDs live at home and are cared for by their relatives (Kasper, 2014). Because the US healthcare system focuses mainly on acute care, relatives provide more than 80% of the long-term care for IWDs (Friedman et al., 2015; Kasper et al., 2015). For these reasons, caregiver support is a key component of the National Alzheimer’s Project Act (U.S. Department of Health & Human Services, 2016).

Most family caregiver interventions have been designed for non-Latino whites and results might not generalize to other groups due to linguistic, cultural, and contextual reasons (Gilmore-Bykovskyi et al., 2018; L. N. Gitlin et al., 2015; Pendergrass et al., 2015). The number of Latino IWDs is projected to increase from 379,000 in 2012 to 3.5 million by 2060, more than any other group (Wu et al., 2016). Latinos are more likely to become family caregivers than non-Latino whites (National Alliance for Caregiving, 2008). Latinos also provide more intense and longer caregiving, and experience higher levels of caregiver depression and burden (Gallagher-Thompson et al., 2003; Garcia, 2000; Hinton et al., 2006; Hinton et al., 2003; Napoles et al., 2010; National Alliance for Caregiving, 2008; Pinquart & Sörensen, 2005; Talamantes & Aranda, 2004). However, despite their high interest in participating in caregiver support interventions (J. Perales et al., 2018), Latino caregivers of IWDs are less likely to use caregiver support services (Monahan et al., 1992; Scharlach et al., 2008). This disparity is partly due to the inconvenience of leaving the IWDs and barriers related to transportation, financial, language and cultural aspects (Monahan et al., 1992; Scharlach et al., 2008). The need for targeted caregiver support interventions among Latinos is therefore crucial. This need is in line with the National Institute on Aging’s call to address health disparities in aging research (National Institute on Aging, 2018).

Text messaging offers distinct advantages over websites and apps for delivering interventions (Grossman et al., 2018; Kajiyama et al., 2018; Waller et al., 2017). While nearly all Latinos engage in text messaging, Latinos’ low use of websites and apps could perpetuate disparities in access to caregiving support (Anderson, 2015). Caregiver interventions for Latinos need to capitalize on text messaging as text message interventions 1) are effective in other health conditions, 2) can be used anywhere at any time, 3) are more cost-effective than other delivery systems, 4) can be personalized to caregivers’ preferences and characteristics including language, culture, and needs, 5) are highly scalable among Latinos, as most own a cellphone with texting capabilities; more than other groups, and 6) have been specifically shown to engage Latinos (Cartujano-Barrera et al., 2020; Guerriero et al., 2013; Hall et al., 2015; Pew Research Center, 2021; Schilling et al., 2013; Zurovac et al., 2012).

To address Latinos’ disparities in access to caregiving support, we developed *CuidaTEXT* (a Spanish play on words for self-care and texting). To our knowledge, this is the first text message intervention for caregiver support of IWDs among Latinos or any other ethnic group. Only one other text message intervention exists in the context of dementia (Lincoln et al., 2019). However, that intervention was designed to increase dementia literacy among non-Latino Black users and is not geared towards Latinos nor caregivers specifically. The aim of this manuscript is to describe the development of *CuidaTEXT*, a text message dementia family caregiver support intervention for Latinos. This development corresponds to Stage 1a of the NIH Stage Model for Behavioral Intervention Development (intervention generation) (Onken et al., 2014). This intervention will be later feasibility-tested (Stage 1b) among Latino family caregivers of 20 IWDs (Clinicaltrials.gov NCT04316104).

## Methods

This was a mixed-methods project guided by user-centered design principles (International Organization for Standardization, 2018). The basis for user-centered design is that gathering and incorporating feedback from users into product design will lead to a more usable and acceptable product. Mixed-methods are required given the lack of literature on text message interventions for Latino family caregivers of IWDs and the strengths of a combined qualitative and quantitative approach (Creswell & Clark, 2017). We followed six user-centered design stages informed by previous research used to develop successful behavioral intervention software (Vilardaga et al., 2018). All study procedures were approved by the Institutional Review Board of the University of Kansas Medical Center (STUDY00144478). All participants gave written informed consent. The user-centered design stages included:

### Stage 1: Selection of design principles

We specified two design principles. First, we chose Social Cognitive Theory as the main behavior change principle (Bandura, 1972). This principle has been used in previous text message interventions successfully (Cartujano-Barrera et al., 2020). The Social Cognitive Theory informs the identification of barriers to desired behaviors, setting of realistic goals, encouragement of gradual practice to achieve performance accomplishments of healthy behaviors (e.g., relaxation techniques or exercising), integration of testimonials and videos to promote vicarious learning, integration of praise to elicit social persuasion, and education to increase dementia knowledge. Second, we chose the Stress Process Framework (Pearlin et al., 1990) to guide the development of messages to encourage coping and social support behaviors (mediators), which are aimed at improving role strains (e.g., perceived income adequacy, family interaction); intrapsychic strains (e.g., mastery, self-esteem, loss of self); and ultimately, outcomes (e.g., caregiver depression, affect, self-perceived health).

### Stage 2: Vendor collaboration for text message design and delivery

This stage aimed to materialize the vision and design specifications of *CuidaTEXT*. We identified a vendor that could provide message personalization and scheduling, use conditional branching logic for text message responses, track information and have an affordable cost. Their services included configuration, account setup, initial onboarding, training guides and videos, two-way text messaging with participants, access to the vendor system and technical support, mobile number support, and bug fixes during the intervention. We contracted with them early in the project to avoid delays (e.g., developing the scope of work, registering as a vendor, contracts, and software programming).

### Stage 3: Evidence-based foundation

Our aim at this stage was to identify core content categories based on prior successful behavioral interventions. We searched specifically for general caregiver support interventions considered evidence-based/informed by the Administration for Community Living (Knowles & Gould, 2019), PubMed literature results using the MeSH terms “Caregivers”, “Dementia” and “Hispanic Americans”, and recommendations on behavioral interventions from the research team. Content categories included dementia education, problem-solving skills training, social network support, care management, and referral to community resources (Alzheimer’s Association, 2021; Baruah et al., 2021; Baumel et al., 2018; Family Caregiver Alliance, 2021; Fitzpatrick et al., 2017; Gallagher-Thompson et al., 2015; L. Gitlin & Piersol, 2014; Gonyea et al., 2014; Grossman et al., 2018; Guerra et al., 2011; Hilgeman et al., 2009; Llanque & Enriquez, 2012; National Academies of Sciences & Medicine, 2016; National Institute on Aging, 2021; Parra-Vidales et al., 2017; Possin et al., 2019; Teri et al., 2003; Waller et al., 2017).

### Stage 4: Advisory board guidance

provided expert opinion to inform the text message intervention based on Latino caregiver needs (J. Perales et al., 2018; J Perales et al., 2017; Reynales-Shigematsu et al., 2017; Vilardaga et al., 2018). We conducted five parallel advisory board meetings with up to 6 Latino caregivers and 16 clinicians and researchers (health professionals), each lasting 60 minutes. We used purposive sampling for the Latino caregivers and quota sampling for the health professionals (including at least one person with expertise in psychiatry, social work, neurology, dementia care interventions, Latino research, text message intervention development or behavioral health). We conducted the caregiver advisory board sessions in Spanish and the professional group sessions in English. We held all sessions via Zoom (https://zoom.us/) from December 2020 to May 2021 and recorded each to facilitate notetaking and analysis. The process for each meeting was similar: the research team showed the groups a step-by-step explanation of the components of the study, asked specific questions pertinent to the phase of the study, and then facilitated open discussion about the project. The research team took detailed notes of all sessions, which were used for further analysis. We organized the notes for qualitative review, using a pragmatic approach, a qualitative description methodology, and thematic analysis methods (Basch, 1987; Miles & Huberman, 1984; Neergaard et al., 2009). We coded the content of the notes using Microsoft Word by identifying codes and themes within the text (Glaser & Strauss, 1967). Two researchers (JPP, MAR) conducted independent reviews of the codes and resolved coding disagreements through discussion and consensus.

### Stage 5: Sketching and prototyping

Based on the previous stages, three researchers (JPP, MAR and PEK) brainstormed a pool of potential text messages in English on a shared spreadsheet, later sorting the messages by topic (initial draft keywords). We edited messages following the ‘Seven Principles of Communication’: completeness, concreteness, courtesy, correctness, clarity, consideration and conciseness (Cutlip, 1952). This theory is popular in business communications and has been used in patient reporting (Aggarwal & Gupta, 2001; Sureka et al., 2018). Bilingual, bicultural members of the research team translated the messages into the primary Spanish dialects represented in the US (Mexican and Caribbean).

In addition to the text message libraries, we developed a reference booklet for participants that summarized the purpose of the intervention and its functions. The booklet is not necessary to use *CuidaTEXT*. However, the research team considered it to be useful for those who want to learn about the intervention faster or increase personal sense of agency. Based on our previous development experience with the Latino community (Cartujano-Barrera et al., 2020; J. Perales et al., 2018), we made the booklet available in both English and Spanish and used lay language and a pictorial format. Seven research team members tested the text message prototype powered by the vendor on their own cellphones from early June to August 2021 and provided feedback that was used for message refinement iteratively as suggested by the literature (Buxton, 2010; Cooper et al., 2007). We recorded the feedback via text message responses within the vendor platform and emails from the research team to the vendor’s programmer. We organized the data (text messages and emails) for qualitative review using a process identical to that described in Stage 4.

### Stage 6: Usability testing

aimed to test a short prototype of the text message intervention and assessments among actual Latino caregivers. We used the vendor’s platform to preview the behavior and opinions of diverse Latino caregivers in a variety of key scenarios (i.e., reading specific messages, using keywords, sending texts, opening links to websites, and videos, and downloading PDF files. The testing sessions were conducted via Zoom in June 2021, lasted approximately 90 minutes per person, and were conducted in English and Spanish, based on participant preferences. We took detailed notes of observations during the usability testing sessions, and participants’ comments at the end.

#### Sample and assessment

We recruited five individuals as suggested by software development cost-benefit analyses (Nielsen & Landauer, 1993). In this framework, the first participant discovers most flaws, and after the fifth user, findings tend to repeat without learning much new. Participants were recruited from three previous projects at the research center using purposive sampling. Eligibility criteria included Spanish or English-speaking individuals who were over the age of 18, identified as Latino, reported providing care for a relative with a clinical or research dementia diagnosis, and with an AD-8 cognitive screening score equal or higher than 2, indicating cognitive impairment (Galvin et al., 2005; Pardo et al., 2013). Participants received $20 prepaid gift cards for completing assessments. We employed three usability evaluation modalities. First, direct observation of task completion (e.g., texting keywords) with the intervention prototype via monitoring of participants text message responses. Second, open-ended interviews of user experience with the different tasks and suggested changes to improve the intervention. Third, caregiver participants completed the System Usability Scale about their experience with the prototype (Sauro, 2011). The System Usability Scale is a valid and reliable 10-item 5-point Likert scale. According to the developers of the scale, scores above 68 out of 100 indicate higher levels of usability. We modified the design features iteratively after each participant and provided the new version to the following participant, as suggested by the literature (Rubin & Chisnell, 2008). After two consecutive participants reviewed and approved a text message, the following participant received a text message with different features. In addition to the usability evaluation, we administered a survey gathering baseline characteristics.

#### Data analysis

We analyzed the qualitative data (detailed notes) from the open-ended interviews using a process identical to that described in Stage 4. We analyzed quantitative baseline characteristics using central tendency estimates, frequencies and percentages on SPSS Software (IBM Corp., 2013).

## Results

This section focuses on Stages 4-6 of the user-centered design proposed in this project. We first summarize the findings from each stage. Second, we explain how these findings informed the intervention development within each stage. Third, we describe the final version of the intervention.

### Findings from advisory board guidance (Stage 4)

Table 1 shows the themes, subthemes, and descriptions of aspects to consider in the development of *CuidaTEXT*, according to the advisory boards. Both Latino caregivers and health professionals contributed to all themes. The advisory board emphasized that messages should include specific content on domains that include logistics (e.g., guidance about *CuidaTEXT’s* functions, motivation messages), social support, caregiver needs, care recipient needs, preparation for the care recipient’s death, and reminders for doctor visits or medicines. Advisory board members highlighted the importance of allowing the inclusion of more than one relative within the family to reduce burden and increase social support. The advisory board suggested that the domains of text messages sent to caregivers should alternate often and be tailored to the needs of caregivers. The preferences of caregiver advisory board members varied widely with respect to how many messages per day *CuidaTEXT* should send participants. One participant emphasized that they would abandon the intervention if they received more than one per day except at the beginning, which required more messaging. Others wanted to receive five or more per day. Eventually a consensus was met that *CuidaTEXT* should tailor the number of messages to the preferences of caregivers. The advisory board emphasized the need to make keyword names as simple and recognizable as possible and suggested several edits in line with this idea. An advisory board member, guided by her experience, highlighted the scarce existing resources for caregivers with hearing issues and suggested that *CuidaTEXT* was made as ‘hearing impairment-friendly’ as possible. After showing the *CuidaTEXT* reference booklet to the advisory board, they suggested several edits to simplify it.

**Table 1.** Themes, subthemes and descriptions of topics elicited during the advisory board sessions with Latino caregivers and health professionals.

| Themes and subthemes | Description |
| --- | --- |
| Messages should include specific content |  |
| Logistics | Motivate participants to be in the program. |
| Dementia education | Address the whole family to be all on the same page; dementia stages and variation between individuals; signs and symptoms; address stigma by normalizing dementia. |
| Social support | Communication with the IWD, include concrete examples (e.g., allowing IWDs enough time to answer); resources (e.g. Alzheimer's Association, health professionals, services in Spanish, support groups, legal assistance); communication with the health provider (e.g. expectations, what to report, encouraging clinical diagnosis, requesting interpreters); improve family communication (e.g. understanding family roles and find strengths, importance of family support, disclosing the diagnosis to the family); develop active listening skills (e.g., reflecting, non-verbal language). |
| Caregiver needs | Cope with depression, anxiety, and stress (e.g., relaxation); choose one's battles; use sense of humor; find positive aspects of caregiving; address loneliness; cope with loneliness; capitalize on spirituality; maintain a healthy lifestyle. |
| Care recipient needs | Address behavioral symptoms as specifically as possible; address aggressive behavior (e.g., understanding the disease is causing it, distraction techniques, communication when the person is aggressive); address anxious behavior (distraction techniques and prevention); help the IWD; address daily care of the IWD including healthy eating, dressing, hygiene, and doing fulfilling activities. |
| Preparation for the care recipients' death | Information about what to expect at the end of the IWD's life; grieving strategies and tips. |
| Appointments for the doctor or medicine | Set notifications to remind caregivers of medications or doctor visits. |
| Need to integrate other family members | More than one family member should be allowed to participate, to: share responsibilities, avoid burdening one caregiver by having to educate the rest, promote collective understanding about what the IWD is experiencing, and reduce the isolation of the caregiving process. |
| Messages should follow a certain order | Alternate content often (e.g., educational, caregiver tips, resources); tailor content to caregivers' needs. |
| Messages should have a certain dose | Limit mandatory message frequency to one per day (in general); tailor frequency and timing to caregivers' preferences. |
| How to integrate COVID-19 into *CuidaTEXT* | Reliable education about caring during COVID-19, include what to do if infected, risks, vaccines, and vaccine locations; resources for those experiencing technological divide. Since changes in COVID-19 evolve quickly, it was decided not to automate messages and send only ad-hoc information as needed. |
| Some keyword names need editing | Use simple and recognizable keyword names if possible; allow platform recognition of alternative spellings and typos; edit specific keywords: STRESS vs RELAX, GRIEF vs ENDOFLIFE or LOSS, BANO vs ORINAR, DUELO vs FINDEVIDA, PACIENTE vs SERQUERIDO. |
| *CuidaTEXT* could benefit people with hearing impairment | Given that text messages are visual, this intervention can be optimized for people with hearing impairment. |
| Reference booklet needs editing | Shorten booklet; include fewer and more realistic examples by including a matrix of the different keyword messages. |

Building upon the evidence-based foundation established in Stage 3, feedback from this stage informed the sketching and prototyping of *CuidaTEXT*. Some examples of the additions informed by this stage include 1) the addition of all the text message content suggested in this stage, 2) enrolling more than one caregiver per IWD, 3) including high priority message content at the beginning of the intervention (e.g., who to contact in case of elder abuse or suicidal thoughts, removing weapons in the home), 4) sending few daily automatic messages per day to all participants (generally one), 5) developing keywords to tailor the message content and frequency to caregiver’s needs for those who want additional messages, 6) excluding COVID-19 in *CuidaTEXT*, but preparing the live coach for potential ad-hoc questions, 7) refining keyword names, 8) accounting for hearing impairment when developing messages by, for example, ensuring that any videos linked to the text messages have closed caption subtitles, and 9) refining the *CuidaTEXT* reference booklet.

### Findings from sketching and prototyping (Stage 5)

Table 2 shows the themes, subthemes, and descriptions of aspects considered in the development of *CuidaTEXT*, based on the feedback from text messages from the team during the testing of the prototype and suggestions provided by the vendor’s programmer. These aspects resulted in several solutions. First, we allowed the platform to recognize common misspelling or alternative spellings for keywords (e.g., for the keyword ‘Behavior’, the platform should also accept behaviour, behaviors, behaviours and behavour). Second, we edited words to eliminate misspellings or replace words that required high literacy levels or specialized knowledge (e.g., glutes vs rear). Third, we used a vendor-owned link shortener to save text message characters, as using a third-party link shortener could lead to phone carriers identifying messages as spam and subsequently blocking them. Fourth, we added code to embed the participant’s first name in text messages to personalize them. Fifth, we adjusted the time of delivery of each daily automatic text message to account for participants’ time zone. Sixth, we requested that the vendor allow keyword libraries to loop back to the first text message after reaching the last one on the list.

**Table 2.**
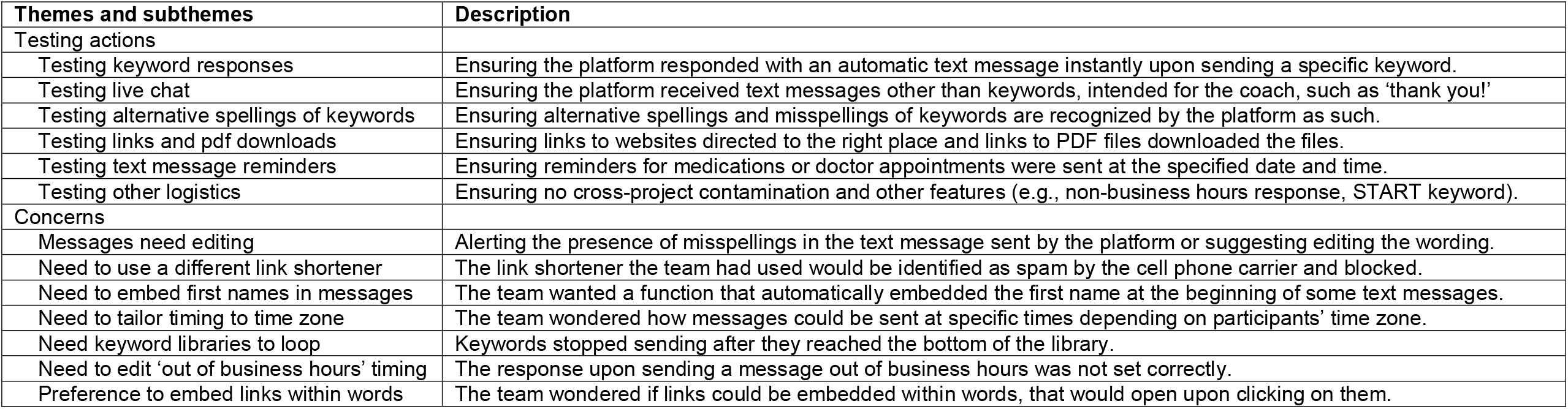
Themes, subthemes and descriptions of topics elicited during the sketching and prototyping stage, gathered via text messages responses from the team and correspondence with the text message vendor.

### Findings from usability testing (Stage 6)

Four of the five participating caregivers in the usability testing were women. The mean age was 44.6 (ranging from 33 to 50). All participants were insured, and their mean level of education was 15.6 (ranging from 12 to 18 years). All participants identified as Latino, two as Native American, one as White and two as more than one race. All participants were born outside the US, including Mexico (n=1), Central (n=2), and South America (n=2). All but one participant completed the intervention and assessments in Spanish and their self-perceived level of spoken English was medium (n=2), high (n=1) and very high (n=2). Participants were daughters (n=3), a son (n=1) and a granddaughter (n=1) of an IWD. Their average care recipient’s age was 77.0 (ranging from 72 to 83).

In general, the participants’ completion of surveys and texting tasks was accomplished without any major issues (e.g., reading specific messages, using keywords, sending texts, opening links to websites and videos, and downloading PDF files). Observations of participants’ reactions during the usability testing and comments at the end of the testing revealed some minor concerns, and generally positive feedback (Table 3). We addressed the concerns in a variety of ways. First, we replaced expressions that were hard to understand (e.g., 24/7 for Spanish-speakers with ‘at any time’). Second, we added context to several text messages to improve understanding. For example, we explained that a ‘Caregiver forum’ is a web platform to share experiences with other caregivers; that the content of a PDF file of a Latin American healthy recipe book alternated pages in English and Spanish; the function of specific keywords; the keyword options using simple graphics; that keywords can be sent more than once for additional messages; and that websites and other resources had a Spanish language option. Third, we tailored the response to the keyword STOP (discontinuing the intervention). We also tailored the notification *CuidaTEXT* automatically sends out when a participant texts outside of business hours by including both languages within the same message, since the platform did not allow separate messages in English and Spanish.

**Table 3.** Themes, subthemes and descriptions of topics elicited during the pilot with five Latino caregiver participants via observation or comments.

| Themes and subthemes | Description |
| --- | --- |
| Participants' concerns |  |
| Expressions are hard to understand | Expressions such as 24/7, or the '+' sign for 'more' were hard to understand, especially for Spanish-speaking Latinos. |
| CuidaTXT is hard to read in Spanish | Spanish-speaking participants had issues pronouncing the original name 'CuidaTXT', and suggested 'CuidaTEXT'. |
| Context is needed for understanding | Some text messages required additional context or an explanation to be understood, for example for the word 'forum'. |
| Branching did not work | In one instance, responding YES or NO to a specific question was not followed by a preconfigured response. |
| Need to edit language of functions | The response upon discontinuing the program or sending a message out of business hours was only in English. |
| Need for caregiver lifestyle messages | One participant suggested adding nutrition as a healthy lifestyle action for caregivers. |
| Other comments from participants |  |
| Satisfaction with the intervention | Satisfaction with the intervention in general, content, logistics and simplicity. |
| Gratitude for the intervention | Expressions of gratitude for developing the intervention. |
| Highlighting need for this intervention | Highlighting the need for this intervention among Latinos and themselves. |

Participants shared mostly positive feedback at the end of the interview, including the following. First, satisfaction with the intervention in terms of general content, logistics, and simplicity was high. Comments included: ‘I think the program is great.’; ‘I love the information and the testimonials.’; ‘The messages made me feel like I’m not alone and put things into perspective.’; and ‘The messages are simple, and the gratitude-theme messages helped.’ Second, participants expressed their gratitude to the *CuidaTEXT* team for developing the intervention. Comments included: ‘Thank you for creating this type of program!’. Third, expressed their need and that of the community to use this intervention. Comments included: ‘I hope we can use it soon because we need it.’; ‘I think it’s going to be very helpful for caregivers emotionally and personally.’ The mean System Usability Scale score was 97 and ranged from 90 to 100, which is above the standard cutoff of 68. These scores indicate the intervention’s usability holds promise.

### Final product

Figure 1 summarizes the final *CuidaTEXT* product, including an example of the three types of text message interaction modalities (daily automatic, keyword-driven and live chat messages) and the reference booklet. The final version of *CuidaTEXT* includes 244 English and 244 Spanish-language messages within the daily automatic text message library. These messages will be automatically sent to all participants starting with approximately three messages per day for the first two weeks, two per day for the following two weeks and one per day for the remainder of the intervention. This daily automatic text message library includes logistics messages that greet the participant upon starting and completing the intervention, explain the intervention functions (e.g., reminding participants the keywords they can use for help with specific topics) and reinforce participants for being in the intervention after two weeks initially and monthly. The remainder of the daily automatic library includes the messages that the research team and advisory board considered to be core from each domain. These domains include messages for 1) dementia education, 2) caregiver self-care messages, 3) support to and from others, 4) education about the dying and grief processes, 5) generic problem-solving strategies for behavioral symptoms, 6) specific strategies to help with the daily care of the IWDs, and 7) specific strategies to help address or cope with the IWDs’ behavioral symptoms.

**Figure 1.**
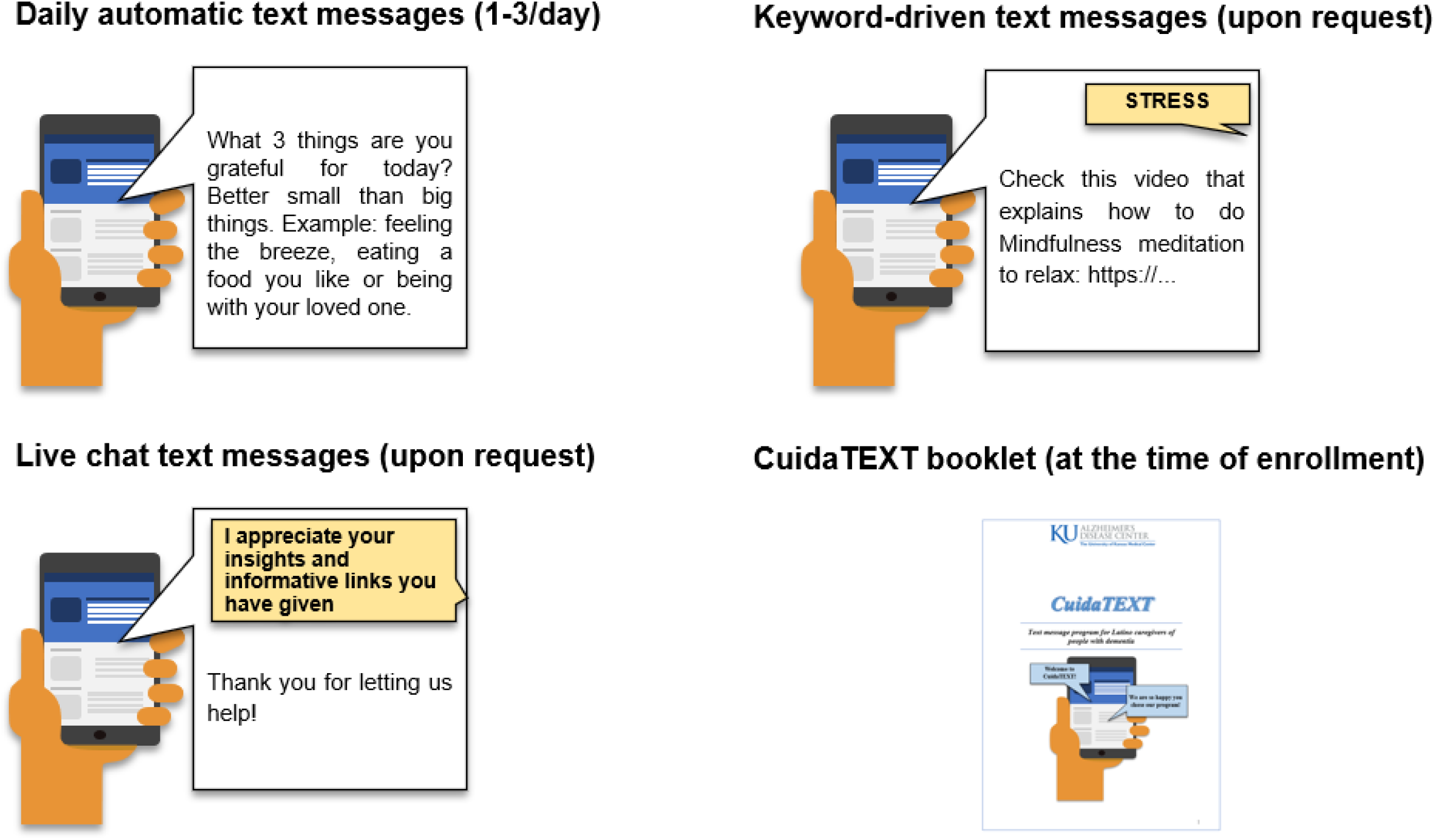
Final *CuidaTEXT* product: Text message interaction modalities and booklet

*CuidaTEXT* also contains messages for two types of keyword-driven messages, the content keywords and menu keywords. Content keywords automatically send tips, resources, or other types of content in response to text messages that include a specific keyword (e.g., STRESS, RESOURCES). These keywords reflect the same domains as the daily automatic text message library, except, their content is not considered to be core, but rather an in-depth expansion for those who need further support with those domains. Menu keywords simply remind the participant which content keywords are in that category. For example, texting the menu keyword ‘CAREGIVER’ will drive an automatic response reminding participants that content keywords within that domain include ‘STRESS’, ‘WELLBEING’ and ‘LIFESYLE’. Table 4 shows the function and an example of each content keyword, the menu keyword they belong in, and the number of messages in each of the content keyword libraries.

**Table 4.**
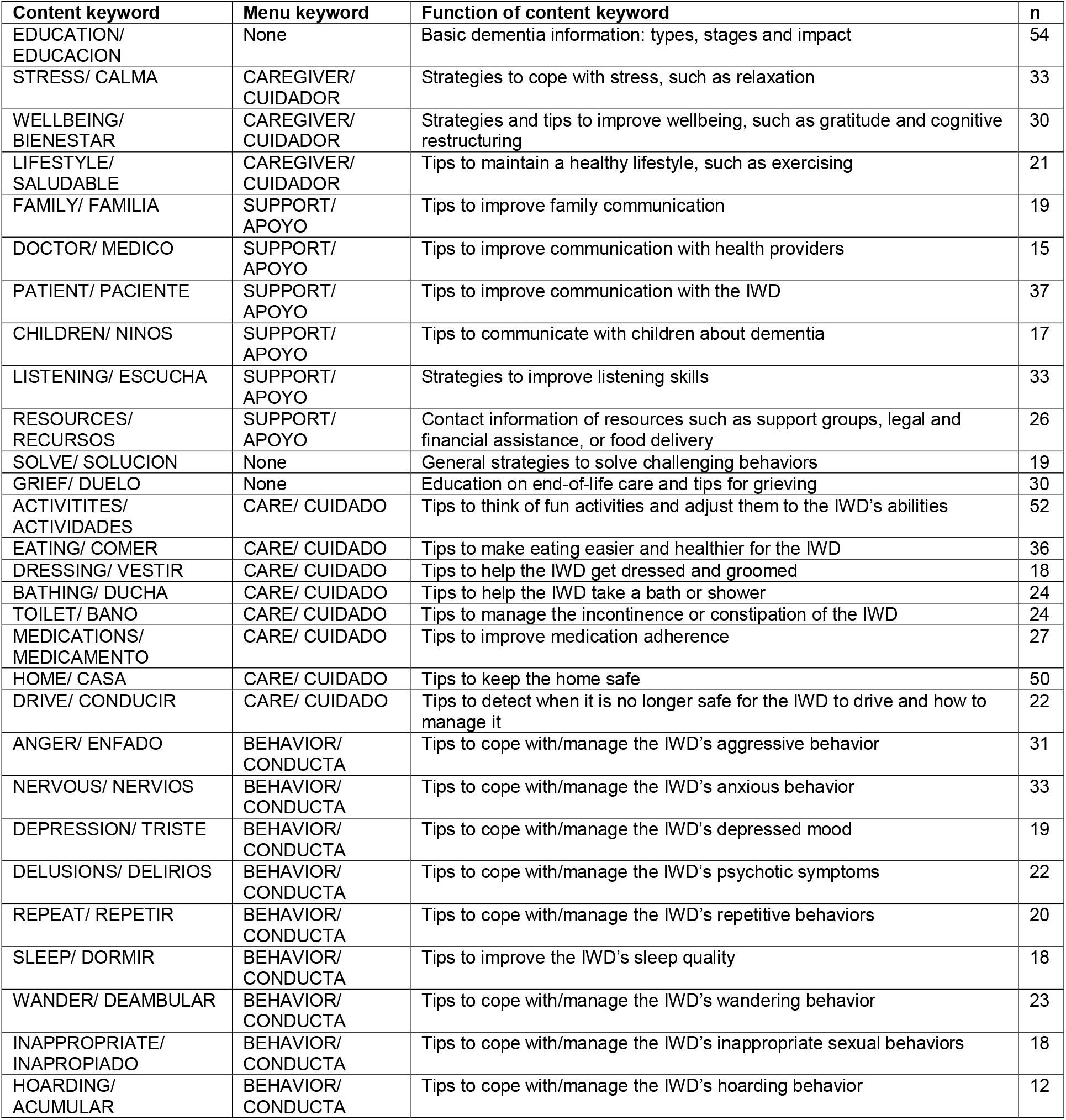
Keywords, their function and size of each keyword library in number of messages (n).

| Content keyword | Menu keyword | Function of content keyword | n |
| --- | --- | --- | --- |
| EDUCATION/<br>EDUCACION | None | Basic dementia information: types, stages and impact | 54 |
| STRESS/ CALMA | CAREGIVER/<br>CUIDADOR | Strategies to cope with stress, such as relaxation | 33 |
| WELLBEING/<br>BIENESTAR | CAREGIVER/<br>CUIDADOR | Strategies and tips to improve wellbeing, such as gratitude and cognitive restructuring | 30 |
| LIFESTYLE/<br>SALUDABLE | CAREGIVER/<br>CUIDADOR | Tips to maintain a healthy lifestyle, such as exercising | 21 |
| FAMILY/ FAMILIA | SUPPORT/<br>APOYO | Tips to improve family communication | 19 |
| DOCTOR/ MEDICO | SUPPORT/<br>APOYO | Tips to improve communication with health providers | 15 |
| PATIENT/ PACIENTE | SUPPORT/<br>APOYO | Tips to improve communication with the IWD | 37 |
| CHILDREN/ NINOS | SUPPORT/<br>APOYO | Tips to communicate with children about dementia | 17 |
| LISTENING/ ESCUCHA | SUPPORT/<br>APOYO | Strategies to improve listening skills | 33 |
| RESOURCES/<br>RECURSOS | SUPPORT/<br>APOYO | Contact information of resources such as support groups, legal and financial assistance, or food delivery | 26 |
| SOLVE/ SOLUCION | None | General strategies to solve challenging behaviors | 19 |
| GRIEF/ DUELO | None | Education on end-of-life care and tips for grieving | 30 |
| ACTIVITIES/<br>ACTIVIDADES | CARE/ CUIDADO | Tips to think of fun activities and adjust them to the IWD's abilities | 52 |
| EATING/ COMER | CARE/ CUIDADO | Tips to make eating easier and healthier for the IWD | 36 |
| DRESSING/ VESTIR | CARE/ CUIDADO | Tips to help the IWD get dressed and groomed | 18 |
| BATHING/ DUCHA | CARE/ CUIDADO | Tips to help the IWD take a bath or shower | 24 |
| TOILET/ BANO | CARE/ CUIDADO | Tips to manage the incontinence or constipation of the IWD | 24 |
| MEDICATIONS/<br>MEDICAMENTO | CARE/ CUIDADO | Tips to improve medication adherence | 27 |
| HOME/ CASA | CARE/ CUIDADO | Tips to keep the home safe | 50 |
| DRIVE/ CONDUCIR | CARE/ CUIDADO | Tips to detect when it is no longer safe for the IWD to drive and how to manage it | 22 |
| ANGER/ ENFADO | BEHAVIOR/<br>CONDUCTA | Tips to cope with/manage the IWD's aggressive behavior | 31 |
| NERVOUS/ NERVIOS | BEHAVIOR/<br>CONDUCTA | Tips to cope with/manage the IWD's anxious behavior | 33 |
| DEPRESSION/ TRISTE | BEHAVIOR/<br>CONDUCTA | Tips to cope with/manage the IWD's depressed mood | 19 |
| DELUSIONS/ DELIRIOS | BEHAVIOR/<br>CONDUCTA | Tips to cope with/manage the IWD's psychotic symptoms | 22 |
| REPEAT/ REPETIR | BEHAVIOR/<br>CONDUCTA | Tips to cope with/manage the IWD's repetitive behaviors | 20 |
| SLEEP/ DORMIR | BEHAVIOR/<br>CONDUCTA | Tips to improve the IWD's sleep quality | 18 |
| WANDER/ DEAMBULAR | BEHAVIOR/<br>CONDUCTA | Tips to cope with/manage the IWD's wandering behavior | 23 |
| INAPPROPRIATE/<br>INAPROPIADO | BEHAVIOR/<br>CONDUCTA | Tips to cope with/manage the IWD's inappropriate sexual behaviors | 18 |
| HOARDING/<br>ACUMULAR | BEHAVIOR/<br>CONDUCTA | Tips to cope with/manage the IWD's hoarding behavior | 12 |

Any text message other than keywords sent by participants will be received as a live chat by a bilingual and culturally proficient coach trained in dementia care. The coach will assist participants in whatever their need is (e.g., additional information about a caregiver grant, programing 3-way calls with a clinic, etc.). Also, the final version of the *CuidaTEXT* reference booklet includes 19 pages with nine chapters: 1) What is dementia, 2) Signs and symptoms of dementia, 3) Why focus on Latino caregivers, 4) *CuidaTEXT* (Automatic Messages), 5) Assistant, 6) Notifications, 7) Keywords, 8) Materials, and 9) Contact information.

## Discussion

This study aimed to describe the development of *CuidaTEXT*, a text message dementia family caregiver support intervention for Latinos. We followed user-centered design principles, to ensure the intervention’s tailoring and usability among Latino caregivers of IWDs. After a series of user-centered design stages, *CuidaTEXT’s* prototype showed a very high usability score, indicating great promise for the intervention’s feasibility and acceptability.

To our knowledge, this is the first text message intervention for caregiver support of IWDs among Latinos and any other ethnic group. Very few evidence-based and culturally-tailored caregiver support interventions have been developed for Latinos. These interventions include fotonovelas, webnovelas, support groups, care management, and psychoeducational programs (Chodosh et al., 2015; Gallagher-Thompson et al., 2015; Gonyea et al., 2014; Kajiyama et al., 2018; Llanque & Enriquez, 2012). The modality of all these interventions has been individual or group face-to-face, computer, telephone or mail based. *CuidaTEXT* has the potential to address implementation gaps in these interventions by: 1) increased accessibility compared to face-to-face or web-based interventions, 2) improved acceptability compared to phone-based interventions, 3) tailoring the content to the needs of caregivers rather than using rigid curriculums, 4) addressing stigma by sending texts privately to the caregivers’ cell phone, and 5) reduced demand on the healthcare workforce to deliver the intervention, therefore improving fidelity and facilitating future scaleup of the intervention. While *CuidaTEXT* was developed for Latinos, similar interventions may be beneficial for other ethnic groups, especially those in rural areas, given the nearly universal cellphone ownership of most populations in the US (Pew Research Center, 2021).

The advisory board suggested text message content related to dementia education, social support, care and caregiver needs, community resources and appointment reminders. These domains are the domains most commonly included in multidomain caregiver support interventions, which have been shown to be more efficacious than single-domain interventions (Grossman et al., 2018; National Academies of Sciences & Medicine, 2016). As mentioned in a recent federally commissioned report, of all interventions to improve caregiver well-being, multicomponent interventions use the most targeted components, and they possibly address at least one critical need across a wide range of individual caregiver needs, thus improving caregiver and IWD outcomes (Butler et al., 2020).

The advisory board encouraged the inclusion of more than one family member per IWD. This idea is in line with the fact that caregiving tasks and decision-making among Latinos are more likely to be shared by multiple relatives of the IWDs (Apesoa-Varano et al., 2015; Gallagher-Thompson et al., 2003). In fact, interventions rarely include other family members, which is likely a reflection of centering interventions on non-Latino White caregivers (Butler et al., 2020; National Academies of Sciences, 2021). According to our advisory board, potential benefits of including more than one family member may include improving caregiving quality and reducing caregiver burden.

This study has several limitations. First, most participants in the usability testing stage identified as the adult children of the IWDs, were women, were relatively highly educated and had at least a medium level of English proficiency. This sample may have placed a higher focus of the refinement of the intervention on these groups rather than men, spousal caregivers, and those with lower educational attainment or limited English proficiency, who may have uniquely different needs. However, most caregivers are women and representatives of many of these other groups were represented in other stages of the development of *CuidaTEXT*. Second, the eligibility to participate in the caregiver advisory board sessions and the usability testing was based on self-report of the care recipient’s dementia status. Third, *CuidaTEXT* does not provide automated content tailoring based on baseline characteristics or tailoring of the timing based on caregiver preferences, as suggested by some advisory board participants. A higher level of automated tailoring might address participants’ needs in more depth and tailoring to the time in which participants receive text messages might lead to higher acceptability. However, we consider that including keyword-driven messages addresses many of the same concerns that automated content and time tailoring address. Keyword-driven messages also reduce the reliance on a baseline assessment. Any need beyond those addressed by the keyword-driven messages can also be addressed via live chat with a coach. Fourth, the current study assessed the usability of the *CuidaTEXT* prototype. While this prototype included the most relevant aspects of *CuidaTEXT*, future studies need to assess the usability of the whole intervention.

This study has implications for public health, clinical practice, and research. Regarding the public health implications, *CuidaTEXT* or similar interventions have high potential for implementation, given their ubiquitous accessibility and reliance on technology rather than human labor. Experts in dementia caregiver interventions highlight the importance of designing interventions with implementation in mind from the beginning of the intervention for its future success (Gaugler et al., 2021). The user-centered design employed to develop this intervention will increase the chances of this intervention to be usable, acceptable, feasible and effective in the future. Regarding clinical practice, usability testing participants described the prototype as something that was needed by them and the Latino community. If *CuidaTEXT* proves to be effective in future studies, this intervention could be easily implemented in clinics and community organizations, by having the caregivers send a text message to enroll or by having staff enter their phone numbers and names on a website. The text message modality may be combined with other modalities to enhance its effectiveness. For example, coaches or social workers could, in addition to interacting via live chat text messages, conduct ad hoc visits or calls with the caregiver. Other findings from this study might also be useful to clinicians, including the need to consider shared caregiver roles within Latino families and other preferences. Findings reported in this manuscript may also inform future research. Future text message studies (whether they are dementia-related or not) might decide to address the content or logistics of their interventions based on the feedback we received from the advisory board sessions or usability testing feedback. Caregiver studies might want to test the efficacy of the same caregiver support intervention to only the primary caregiver vs multiple caregivers within the same family. Future studies will test the feasibility and acceptability of *CuidaTEXT* among Latino caregivers, including those with hearing impairment. If successful, we will conduct a fully powered randomized control trial to assess its efficacy.

## Conclusion

This study describes the development of *CuidaTEXT*, the first tailored text message intervention specifically designed to support dementia family caregivers in the Latino community. The prototype of *CuidaTEXT* has shown very high usability, addresses Latino caregiver needs and has potential for widespread implementation. Findings from several stages of the user-centered design provide useful information to guide the development and refinement of caregiver support interventions for Latinos and other groups, contributing to efforts to address dementia disparities among Latinos and gaps in the implementation of caregiver support interventions for this sizable population. We will soon test the feasibility and acceptability of this promising intervention (*CuidaTEXT*) in a one-arm trial with Latino family caregivers of 20 IWDs (Clinicaltrials.gov NCT04316104).

## Clinical implications

- Latino family caregivers of individuals with dementia face many barriers to caregiver support access that may be alleviated through culturally tailored text message interventions.
- The prototype of *CuidaTEXT*, a text message intervention for family caregiver support has high usability and addresses Latino caregivers’ needs.

## Data Availability

Data produced in the present study are available upon reasonable request to the authors.

## Author note

### Funding

This work was supported by the NIH under Grants R21 AG065755, K01 MD014177, and P30 AG072973.

## Acknowledgements

Dr. Perales-Puchalt thanks the national and local organizations that have partnered with him to conduct this and other research since 2015. Dr. Perales also thanks research participants included in all stages of this research as well as anyone who has contributed directly and indirectly to this research.

## Data availability statement

None

## Biographical note

Jaime Perales-Puchalt, PhD, MPH: Dr. Perales-Puchalt is an Assistant Professor at the University of Kansas Alzheimer’s Disease Research Center. His background is in psychology and public health. In addition to conducting research in Spain, England and the United States, Dr. Perales Puchalt has collaborated with teams from other countries in the European Union and the Americas. With a primary focus on reducing Latino dementia disparities, his research has led to the development of a dementia educational/recruitment tool for Latinos. He has also studied the risk of dementia and mild cognitive impairment among sexual and ethnoracial minorities. He currently leads federally funded grants to understand and reduce disparities in dementia care among Latinos. One of those grants includes the first text message caregiver support intervention for Latino family caregivers (*CuidaTEXT*).

Mariola Acosta-Rullán, MS: Mariola Acosta Rullán is a doctoral student in gerontology at the University of Southern California. She has a master’s degree in clinical psychology. Acosta Rullán was selected to be part of the Kansas Dementia & Aging Research Training Internship Program at the KU Alzheimer’s Disease Center, mentored by Dr. Perales-Puchalt. She is interested in dementia care and disparities among ethnic minorities due to the prevalence of Alzheimer’s disease in Hispanics and Latinos.

Mariana Ramírez-Mantilla, MSW: Mariana Ramirez is the Director of Juntos; Center for Advancing Latino Health, at the University of Kansas Medical Center. She works leading core administrative functions at Juntos including recruitment, implementation, and evaluation of several research studies. Ms. Ramírez is actively involved in several community boards across the state of Kansas.

Paul Espinoza-Kissell, BS: Paul Espinoza Kissell is a doctoral student in Health Services Research, Policy, and Administration at the University of Minnesota. He completed a post-baccalaureate research education program at the University of Kansas in 2021. Espinoza Kissell graduated from University of Wisconsin-Milwaukee with a bachelor’s degree in psychology. He is also developing and implementing an independent research project with KU Medical Center faculty mentor Dr. Perales-Puchalt.

Eric D. Vidoni, PhD: Eric Vidoni, PT, PhD, is Director of the Outreach, Recruitment and Education Core for the University of Kansas Alzheimer’s Disease Research Center and an Associate Professor in the Department of Neurology at the University of Kansas Medical Center. Dr. Vidoni got his Bachelor’s of Science in Kinesiology from the University of Illinois, Urbana-Champaign in 2001. In 2008, he completed his MSPT and PhD as part of a joint curriculum at the University of Kansas. He supports and directs the Kansas Dementia and Aging Research Training Program. As core director, he organizes dementia-focused educational outreach opportunities for healthcare providers and the lay public throughout the region.

Michelle Niedens, LSCSW: Michelle Niedens, is director of the Cognitive Care Network, a community-based program focused on early detection, provider partnerships, and integration of interdisciplinary supports. Ms. Niedens’ background is in social work. Ms. Niedens’ special interests include assessment and intervention for associated neuropsychiatric challenges and advancing programs and services for individuals living with early-stage dementia.

Edward F. Ellerbeck, MD: Dr. Ellerbeck is a Professor of Population Health and the Director of the Clinical and Translational Research Education Center at the University of Kansas Medical Center. He has over 25 years of experience in measuring and improving the quality of medical care, particularly in underserved rural and minority communities. His area of interest is in the development of system changes to enhance prevention and treatment of chronic diseases.

Ladson Hinton, MD: Dr. Hinton is a Professor at UC Davis. He is a geriatric psychiatrist, clinical and services researcher, and social scientist. Over the past two decades, he has conducted interdisciplinary research to better understand the cultural and social dimensions of late life depression and dementia-related illness and caregiving experience among older adults and their families. He has applied this knowledge to develop innovative and culturally appropriate intervention approaches to overcome gaps and disparities in healthcare.

Linda Loera, MA: Linda Loera is a Program and Education Manager with the Alzheimer’s Association, CA Southland Chapter. Her responsibilities include educating the community about Alzheimer’s disease, participating in community events to bring awareness to the disease, and supporting families and caregivers through care consultations, support groups and providing information and resources.

A. Susana Ramírez, PhD, MPH: Dr. Ramírez is an Associate Professor of Public Health Communication at the University of California in Merced. As an infodemiologist, Dr. Ramírez applies communication science to advance public health goals for rural and Latino populations.

Esther Lara, MSW: Ms. Lara is a Clinical Social Worker for the UC Davis Alzheimer’s disease center. Ms. Lara’s special interests are in furthering research opportunities for the Latino community in the areas of dementia care, treatment, and caregiving.

Amber Watts, PhD: Dr. Watts is an Associate Professor of Clinical Psychology at University of Kansas and Research Fellow of the KU Alzheimer’s Disease Center. Her doctoral degree is in Gerontology and she conducts biopsychosocial research and behavioral interventions on preventable risk factors for cognitive decline and Alzheimer’s disease.

Kristine Williams, RN, PhD, FNP-BC, FGSA, FAAN E: Dr. Williams is a nurse gerontologist and the E. Jean Hill Professor at the University of Kansas School of Nursing. Her research tests interventions designed to improve the quality of life of people with ADRD and their families.

Jason Resendez, BA: Mr. Resendez Jason is the Executive Director of the UsAgainstAlzheimer’s Center for Brain Health Equity and head of the LatinosAgainstAlzheimer’s Coalition. From clinical trial inclusion to paid family leave for dementia caregivers, he champions brain health equity at every level of the healthcare system. Mr. Resendez has contributed to peer-reviewed research on the socioeconomic impacts of brain health inequities and on the science of community engagement in neuroscience. He is currently the principal investigator of a CDC Healthy Brain Initiative cooperative agreement.

Jeffrey Burns, MD: Dr. Burns is the Director of the KU Alzheimer’s Disease Center and Professor of Neurology. Dr. Burns also runs the center’s Clinical Core. He has been appointed to the Kansas Governor’s Work Group to establish a statewide plan for addressing dementia. Dr. Burns has key expertise in directing dementia care networks and in treating individuals with dementia.

